# Trends in cardiovascular disease risk factors by body mass index category among adults in England 2003-18: analysis of repeated cross-sectional national health surveys

**DOI:** 10.1101/2020.09.02.20186619

**Authors:** Shaun Scholes, Linda Ng Fat, Jennifer S Mindell

**Author notes:** Address correspondence to: Shaun Scholes, PhD, Department of Epidemiology and Public Health, University College London, 1-19 Torrington Place, London, WC1E 6BT, United Kingdom.

## Abstract

**Objective:** Favourable trends in cardiovascular disease (CVD) risk factors at the population level potentially mask differences within high- and low-risk groups. Data from annual, repeated cross-sectional surveys (Health Survey for England 2003-18) was used to examine trends in the prevalence of key CVD risk factors by body mass index (BMI) category among adults aged 16 years or older (n = 115,860).

**Methods:** Six risk factors were investigated: (i) current cigarette smoking; (ii) physical inactivity (< 30 minutes of moderate-to-vigorous physical activity per week); (iii) drinking above recommended daily alcohol limits; (iv) hypertension (measured blood pressure ≥140/90mmHg or use of medicine prescribed for high blood pressure); (v) total diabetes (reported diagnosed or elevated glycated haemoglobin); and (vi) raised total cholesterol (≥5mmol/L). Age-standardised risk factor prevalence was computed in each four-year time period (2003-06; 2007-10; 2011-14; 2015-18) in all adults and by BMI category (normal-weight; overweight; obesity). Change in risk factor prevalence on the absolute scale was computed as the difference between the first and last time-periods, expressed in percentage points (PP).

**Results:** Risk factor change varied by BMI category in a number of cases. Current smoking prevalence fell more sharply for normal-weight men (−8.1 PP; 95% CI: -10.3, -5.8) versus men with obesity (−3.8 PP; 95% CI: -6.2, -1.4). Hypertension remained at a stable level among normal-weight men but decreased among men with obesity (−4.1 PP; 95% CI: -7.1, -1.0). Total diabetes remained at a stable level among normal-weight adults, but increased among adults with obesity (men: 3.5 PP; 95% CI: 1.2, 5.7; women: 3.6 PP; 95% CI: 1.8, 5.4). Raised total cholesterol decreased in all BMI groups, but fell more sharply among women with obesity (−21 PP; 95% CI: -25, -17) versus their normal-weight counterparts (−16 PP; 95% CI: -18, -14).

**Conclusions:** Relative to adults with normal weight, greater reductions in hypertension and raised total cholesterol among adults with overweight and obesity reflect at least in part improvements in screening, treatment and control among those at highest cardiovascular risk. Higher levels of risk factor prevalence among adults with overweight and obesity, in parallel with rising diabetes, highlight the importance of national prevention efforts to combat the public health impact of excess adiposity.

## Introduction

A large body of individual-level epidemiologic studies have documented the higher rates of cardiovascular disease (CVD) outcomes associated independently with modifiable risk factors (1, 2). For example, excess body weight accounted for approximately four million deaths worldwide in 2015; CVD accounted for nearly 70% of deaths related to high body mass index (BMI), of which more than 60% occurred among persons with obesity (3). At the population level, halting the rise of diabetes and obesity, and reducing levels of other CVD risk factors such as current tobacco use, harmful use of alcohol, insufficient physical activity (PA), intake of salt/sodium, and high blood pressure (BP) is a major World Health Organization (WHO) global target for reducing overall mortality from the four main noncommunicable diseases (NCDs) by 25% in 2025 relative to 2010 levels (4).

Obesity prevalence among adults has markedly increased in England over the past 25 years, rising from 16% in 1994 to 28% in 2018 (5), with contributory factors including increases in the availability and affordability of energy dense foods (3) and environmental barriers to PA. At the same time, prevention efforts via greater chronic disease management may have led to sub-groups of the population at higher risk being screened and tested more frequently (6), potentially improving CVD risk factor profiles through lifestyle advice and/or pharmacological treatment of high levels of BP, cholesterol and blood glucose.

Monitoring equity in risk factor reduction requires establishing whether any favourable trends at the population level have been achieved equally within high- and low-risk subgroups, including BMI groups (6–8). Risk factor trends at the population level in England has been investigated previously, including studies that assessed secular changes at the upper tail of the BMI distribution (9). Stability or favourable/unfavourable change in the prevalence of risk factors at the population level potentially masks divergent trends by BMI group. To date, however, no studies have examined trends in both the prevalence and management of CVD risk factors by BMI category in England. Using data from annual, repeated cross-sectional surveys of adults spanning 16 years (Health Survey for England 2003–18), we examined change over time in the prevalence of six key CVD risk factors (4) – smoking, physical inactivity, harmful alcohol consumption, hypertension, diabetes and raised total cholesterol – by BMI category.

## Methods

### Health Survey for England

The Health Survey for England (HSE) is a series of annual surveys designed to measure health and health-related behaviours. Details of the survey methods have been published elsewhere (10). Briefly, new, nationally-representative samples of people living in private households were drawn annually using multistage stratified probability sampling. All adults (aged 16 years or older) at each selected household were eligible. Data were collected at two home visits. First, an interviewer administered a questionnaire on socio-demographic variables, lifestyle behaviours, general health, and self-reported morbidity, and measured height and weight. Secondly, a nurse visited and asked further questions, including current prescribed medication, and collected BP and additional anthropometric measurements and non-fasting blood samples.

Data on height, weight, cigarette smoking and alcohol consumption were collected annually. BP collection took place annually except for 2004 (except for minority ethnic groups); blood samples for cholesterol were taken in 2003, 2006, and each year from 2011 onwards. Information on self-reported diagnosed diabetes and the measurement of glycated haemoglobin (as a marker for undiagnosed diabetes) was collected in 2006, and in each year from 2009 onwards. For the present study we used comparable self-reported physical activity data collected in 2008, 2012, and 2016; alcohol data from 2007 onwards was used to account for the new set of questions on wine consumption (11). Response rates declined over the study-period; estimated response rates were 66% in 2003 and 54% in 2018 (interview); 77% in 2003 and 51% in 2018 (nurse-visit); and 58% in 2003 and 38% in 2018 (blood-samples). Ethical approval was obtained from an NHS Research Ethics Committee prior to starting each year’s survey. Each participant gave verbal consent to be interviewed, visited by a nurse, and have BP and anthropometric measurements taken, and written consent for blood sampling. No specific approval was required for the present analyses of anonymised data.

## Definitions of key variables

### BMI and other CVD risk factors

Height and weight measurements by trained interviewers were taken using standardised protocols. Height was measured using a portable stadiometer with a sliding head plate, a base plate and connecting rods marked with a measuring scale. One measurement was taken with the head positioned in the Frankfort plane. Digital scales were used for weight measurement. A single measurement was recorded: participants who were pregnant, unable to stand, or unsteady on their feet were not weighed. BMI was calculated as weight in kilogrammes divided by height in metres squared (kg/m^2^), and the WHO classification was used to group participants into four mutually exclusive categories: underweight (< 18.5kg/m^2^); normal-weight (18.5–24.9kg/m^2^); overweight (25.0–29.9kg/m^2^); and obesity (≥30.0kg/m^2^) (12). In addition, we present separate estimates for class I obesity (30.0–34.9kg/m^2^) and classes II and III combined (hereafter referred to as class II-III obesity: ≥35.0kg/m^2^).

All risk factors were dichotomised. Self-reported cigarette smoking status was categorised as non- (never smokers, ex-regular smokers) and current smokers. Participants were classified as physically inactive if they spent less than 30 ‘equivalent’ minutes a week engaged in moderate-to-vigorous PA (MVPA), where 1 minute of vigorous PA was equivalent to 2 minutes of moderate PA: details of the PA questionnaire used in the HSE series are described elsewhere (13). Participants who had consumed alcohol in the last week were asked questions on the amounts of different types of alcohol drunk on the day they drank most. For the purposes of the present study, excess drinking was defined as drinking over the recommended daily limits on the heaviest drinking day (4 and 3 units for men and women, respectively) (11). New recommendations introduced in 2016 replaced daily limits with a recommended weekly limit; the threshold based on daily limits was chosen to make use of routinely collected information on the heaviest drinking day.

At the nurse visit, three BP readings were taken from each participant in a seated position at 1-minute intervals with use of an appropriately sized cuff after a 5-minute rest, following a standardised protocol using an Omron digital monitor (Omron HEM-907, Omron Healthcare Co Ltd, Kyoto, Japan). Participants who had exercised, eaten, drunk alcohol, or smoked in the 30 minutes before measurements were excluded from analyses. The mean of the second and third BP readings were used. Survey defined hypertension was defined as systolic blood pressure (SBP) ≥140mmHg, diastolic blood pressure (DBP) ≥90mmHg, or reported taking medication prescribed for high BP. We defined three indicators of hypertension management – diagnosed, treated and controlled – using the subset of participants classified as hypertensive as the denominator. Diagnosed hypertension was defined as a self-report of having been diagnosed as having high BP by a doctor or nurse (only a random subsample of persons aged 65 years or over was asked this question in HSE 2006, requiring use of a specific weight). Treated hypertension was defined as a self-report of taking prescribed medication for high BP. Controlled hypertension was defined as having BP levels below recommended target levels (≤140/90mmHg) (14).

Participants who reported that their doctor had diagnosed them as having diabetes were classified as having doctor-diagnosed diabetes. Glycated haemoglobin (HbA_1c_) was measured from EDTA-blood samples and determined by high-performance liquid chromatography using an automated analyser. Those not reporting diagnosed diabetes were classified as having undiagnosed diabetes if HbA_1c_ was ≥6.5% (prior to HSE 2012) or ≥48mmol/mol (HSE 2012–18). Total diabetes was having doctor-diagnosed or undiagnosed (15). Blood samples were taken for serum total and high-density lipoprotein (HDL) cholesterol. Raised total cholesterol was defined as ≥5 millimoles per litre (mmol/L) regardless of lipid lowering medication use, reflecting National Institute of Health and Clinical Excellence (NICE) guidelines (16). Adjustments to the measured values of HbA_1c_ (from the final quarter of HSE 2013 onwards) and total cholesterol (HSE 2011–16) were applied to account for changes in laboratory equipment.

### Statistical analysis

Analyses were limited to participants aged 16 years or older (n = 137,645) with valid height and weight data (n = 115,860). All analyses were based on complete cases: analytic sample sizes were as follows: current smoking (n = 115,472); physical inactivity (n = 26,051); drinking above recommended daily alcohol limits (n = 83,969); hypertension (n = 71,948); total diabetes (n = 47,818); and raised total cholesterol (n = 43,340).

We chose a-priori to stratify analyses by gender. Available data from four consecutive annual surveys was aggregated into four non-overlapping survey periods (2003–06; 2007–10; 2011–14 and 2015–18) to boost sample sizes and thereby increase the precision of estimates. Risk factor prevalence was estimated by time-period for all adults (i.e. all BMI groups combined) and by BMI category (normal-weight; overweight; obesity; class I obesity; class II-III obesity); estimates are not presented for the underweight category due to the low prevalence (2%). Estimates were age-standardised using the direct method (pooled HSE data as the standard population). The absolute change in prevalence was computed as the difference between the first and last time-periods, expressed in percentage points (PP). Wald tests were used to test the null hypothesis of no change in prevalence between the two estimates. The same procedure was used to compare the difference in prevalence between the first and last time-periods in the overweight and obesity categories versus the normal-weight group. Analyses were repeated on the subset of participants with survey-defined hypertension (n = 23,216) to estimate the change in prevalence of diagnosed, treated and controlled hypertension. Sample sizes were too small to estimate the change in levels of diagnosed diabetes among participants with total diabetes. Changes in diagnosed and undiagnosed diabetes by BMI category was therefore estimated using all adults as the denominator.

All analyses accounted for the complex survey design, incorporating the appropriate weights which accounted for individual nonparticipation to each stage (interview, nurse-visit, blood sample collection) and the geographical clustering of participants in primary sampling units. Statistical significance was set at *p* < 0.05 for two-tailed tests, with no adjustment for multiple comparisons. 95% confidence intervals (95% CIs) are used to convey precision. Dataset preparation and analysis was performed in SPSS v24.0 (SPSS IBM Inc., Chicago, Illinois, USA) and in Stata v15.1 (StataCorp, College Station, Texas, USA), respectively. HSE datasets, including the most recent survey (17), are available via the UK Data Service (http://www.ukdataservice.ac.uk); statistical code to enable replication of our results (using the datasets deposited at the UKDS) is available on request from the corresponding author.

## Results

Table 1 shows the (unweighted) characteristics of the analytical sample (n = 115,860 participants aged 16 or over with valid height and weight measurements) by four-year survey-period. The proportion of participants aged 75 years or over increased from 8.0% in 2003–06 to 10.4% in 2015–18; the proportion with a degree or higher qualification increased from 18.3% to 28.6%. BMI increased on average by 0.6kg/m^2^, reflecting an increase in mean weight of 2kg.

**TABLE 1.** Characteristics of the analytical sample by four-year survey period.

| **Characteristics** | **All**<br>**n (%)** | **2003-06**<br>**n (%)** | **2007-10**<br>**n (%)** | **2011-14**<br>**n (%)** | **2015-18**<br>**n (%)** |
| --- | --- | --- | --- | --- | --- |
| Sample size | 115,860 (100) | 31,421 (100) | 29,708 (100) | 28,204 (100) | 26,527 (100) |
| Male | 52,667 (45) | 14,418 (46) | 13,558 (46) | 12,772 (45) | 11,919 (45) |
| **Age-group:** |  |  |  |  |  |
| 16-34 years | 28,802 (25) | 8222 (26) | 7555 (25) | 6886 (24) | 6139 (23) |
| 35-54 years | 41,273 (36) | 11,541 (37) | 10,626 (36) | 10,051 (36) | 9055 (34) |
| 55-74 years | 35,138 (30) | 9148 (29) | 8838 (30) | 8565 (30) | 8587 (32) |
| 75+ years | 10,647 (9) | 2510 (8) | 2689 (9) | 2702 (10) | 2746 (10) |
| Degree or equivalent | 26,496 (23) | 5747 (18) | 6110 (21) | 7071 (25) | 7568 (29) |
| White | 104,262 (90) | 28,894 (92) | 26,915 (91) | 25,130 (89) | 23,323 (88) |
| BMI (kg/m<sup>2</sup>), mean (sd) | 27.4 (5.4) | 27.1 (5.1) | 27.3 (5.3) | 27.4 (5.4) | 27.7 (5.7) |
| Weight (kg), mean (sd) | 77.0 (17.0) | 76.1 (16.3) | 76.8 (16.7) | 77.3 (17.1) | 78.1 (17.9) |
| Height (m), mean (sd) | 1.7 (0.1) | 1.7 (0.1) | 1.7 (0.1) | 1.7 (0.1) | 1.7 (0.1) |
| **BMI category:** |  |  |  |  |  |
| Underweight | 1776 (2) | 472 (2) | 450 (2) | 428 (2) | 426 (2) |
| Normal-weight | 40,339 (35) | 11,374 (36) | 10,423 (35) | 9712 (34) | 8830 (33) |
| Overweight | 43,526 (38) | 12,014 (38) | 11,205 (38) | 10,651 (38) | 9656 (36) |
| Obesity | 30,219 (26) | 7561 (24) | 7630 (26) | 7413 (26) | 7615 (29) |
| Class I | 20,373 (18) | 5228 (17) | 5222 (18) | 4938 (18) | 4925 (19) |
| Class II-III | 9846 (9) | 2273 (7) | 2408 (8) | 2475 (9) | 2690 (10) |

*Notes:* Analytical sample is based on participants with valid height and weight data. BMI: body mass index; kg: kilogrammes; m: metres; sd: standard deviation. Normal-weight: BMI 18.5 to <25kg/m<sup>2</sup>; overweight: BMI 25 to <30kg/m<sup>2</sup>; obesity: BMI ≥30kg/m<sup>2</sup>; class I obesity 30 to <34.9kg/m<sup>2</sup>; class II-III obesity BMI ≥35kg/m<sup>2</sup>. Estimates are unweighted.

Estimates of CVD risk factor prevalence by survey period and BMI category are shown in Tables 2 and 3 for men and women, respectively. Figure 1 shows the absolute change in prevalence between the first and last time-periods, expressed in percentage points (PP).

**TABLE 2.** Age-standardised risk factor prevalence by BMI group and four-year survey period in men.

| **Risk factor by BMI group** | **2003-06** | **2007-10** | **2011-14** | **2015-18** | **PP change** | **P-value** | **PP change versus normal-weight** | **P-value** |
| --- | --- | --- | --- | --- | --- | --- | --- | --- |
|  | **% (95% CI)** | **% (95% CI)** | **% (95% CI)** | **% (95% CI)** | **% (95% CI)** |  | **% (95% CI)** |  |
| **Current smoking** |  |  |  |  |  |  |  |  |
| All | 25.5 (24.7, 26.3) | 23.1 (22.3, 23.9) | 22.4 (21.5, 23.3) | 19.0 (18.1, 19.9) | -6.5 (-7.7, -5.3) | <0.001 | --- | --- |
| Normal-weight | 32.2 (30.7, 33.7) | 29.1 (27.6, 30.7) | 27.2 (25.6, 28.8) | 24.2 (22.4, 25.9) | -8.1 (-10.3, -5.8) | <0.001 | --- | --- |
| Overweight | 24.6 (23.4, 25.8) | 20.6 (19.5, 21.8) | 21.1 (19.8, 22.4) | 17.6 (16.2, 19.0) | -7.0 (-8.9, -5.2) | <0.001 | 1.0 (-1.8, 3.8) | 0.481 |
| Obesity | 20.6 (18.9, 22.2) | 21.2 (19.4, 23.0) | 21.2 (19.4, 23.1) | 16.7 (15.0, 18.5) | -3.8 (-6.2, -1.4) | 0.002 | 4.2 (1.0, 7.4) | 0.010 |
| Class I | 21.5 (19.5, 23.5) | 21.8 (19.6, 23.9) | 21.4 (19.2, 23.6) | 16.9 (14.9, 19.0) | -4.6 (-7.5, -1.7) | 0.002 | 3.5 (-0.1, 7.1) | 0.055 |
| Class II-III | 18.2 (15.1, 21.2) | 19.3 (16.0, 22.6) | 20.8 (17.6, 24.0) | 16.4 (13.4, 19.4) | -1.8 (-6.1, 2.5) | 0.407 | 6.3 (1.4, 11.1) | 0.011 |
| **Inactivity** |  |  |  |  |  |  |  |  |
| All | --- | 18.2 (17.1, 19.2) | 18.1 (16.6, 19.6) | 18.6 (17.0, 20.1) | 0.4 (-1.5, 2.3) | 0.658 | --- | --- |
| Normal-weight | --- | 17.0 (15.0, 18.9) | 17.3 (14.4, 20.1) | 16.9 (14.1, 19.7) | -0.1 (-3.5, 3.4) | 0.967 | --- | --- |
| Overweight | --- | 16.3 (14.7, 17.9) | 16.0 (14.0, 18.1) | 17.4 (14.8, 19.9) | 1.1 (-1.9, 4.0) | 0.487 | 1.1 (-3.4, 5.6) | 0.621 |
| Obesity | --- | 22.8 (20.3, 25.2) | 22.1 (19.0, 25.1) | 23.7 (20.0, 27.4) | 0.9 (-3.5, 5.4) | 0.682 | 1.0 (-4.5, 6.5) | 0.722 |
| Class I | --- | 21.7 (18.8, 24.7) | 19.5 (16.3, 22.6) | 19.9 (16.6, 23.1) | -1.9 (-6.3, 2.5) | 0.402 | -1.8 (-7.3, 3.7) | 0.518 |
| Class II-III | --- | 26.0 (21.9, 30.1) | 27.9 (21.5, 34.3) | 33.5 (25.3, 41.7) | 7.5 (-1.7, 16.6) | 0.111 | 7.5 (-2.3, 17.3) | 0.132 |
| **Alcohol above daily limits** |  |  |  |  |  |  |  |  |
| All | --- | 42.2 (41.2, 43.2) | 38.4 (37.4, 39.4) | 35.5 (34.4, 36.5) | -6.8 (-8.2, -5.3) | <0.001 | --- | --- |
| Normal-weight | --- | 39.0 (37.3, 40.6) | 35.7 (34.0, 37.4) | 33.8 (32.0, 35.6) | -5.2 (-7.6, -2.7) | <0.001 | --- | --- |
| Overweight | --- | 44.4 (42.9, 45.9) | 40.1 (38.5, 41.6) | 37.2 (35.6, 38.8) | -7.2 (-9.4, -5.0) | <0.001 | -2.0 (-5.2, 1.1) | 0.205 |
| Obesity | --- | 42.2 (40.2, 44.2) | 38.7 (36.7, 40.8) | 34.5 (32.4, 36.6) | -7.7 (-10.6, -4.7) | <0.001 | -2.5 (-6.2, 1.2) | 0.188 |
| Class I | --- | 43.3 (41.0, 45.6) | 40.1 (37.6, 42.7) | 36.5 (33.9, 39.0) | -6.8 (-10.3, -3.4) | <0.001 | -1.7 (-5.8, 2.4) | 0.425 |
| Class II-III | --- | 38.7 (34.7, 42.8) | 35.8 (32.1, 39.5) | 29.4 (25.6, 33.2) | -9.3 (-14.9, -3.8) | 0.001 | -4.2 (-10.2, 1.9) | 0.176 |
| **Hypertension** |  |  |  |  |  |  |  |  |
| All | 31.2 (30.3, 32.1) | 31.5 (30.6, 32.4) | 30.4 (29.4, 31.5) | 28.1 (27.1, 29.2) | -3.0 (-4.4, -1.7) | <0.001 | --- | --- |
| Normal-weight | 21.2 (19.7, 22.7) | 23.2 (21.6, 24.9) | 19.6 (18.0, 21.3) | 19.7 (17.9, 21.5) | -1.5 (-3.9, 0.8) | 0.199 | --- | --- |
| Overweight | 30.5 (29.1, 31.9) | 29.8 (28.4, 31.1) | 30.3 (28.7, 31.9) | 26.0 (24.4, 27.5) | -4.5 (-6.6, -2.5) | <0.001 | -3.0 (-6.1, 0.1) | 0.056 |
| Obesity | 43.5 (41.4, 45.5) | 42.9 (40.7, 45.2) | 42.7 (40.2, 45.1) | 39.4 (37.1, 41.6) | -4.1 (-7.1, -1.0) | 0.009 | -2.5 (-6.3, 1.2) | 0.187 |
| Class I | 42.4 (40.0, 44.8) | 40.3 (37.8, 42.8) | 40.4 (37.6, 43.3) | 35.8 (33.2, 38.4) | -6.6 (-10.1, -3.1) | <0.001 | -5.0 (-9.2, -0.9) | 0.016 |
| Class II-III | 47.2 (43.2, 51.2) | 50.8 (46.4, 55.2) | 48.5 (43.8, 53.3) | 48.4 (43.4, 53.5) | 1.2 (-5.2, 7.6) | 0.712 | 2.7 (-4.1, 9.5) | 0.428 |
| **Raised total cholesterol** |  |  |  |  |  |  |  |  |
| All | 62.0 (60.7, 63.2) | --- | 51.1 (49.8, 52.4) | 45.7 (44.3, 47.0) | -16 (-18, -14) | <0.001 | --- |  |
| Normal-weight | 55.8 (53.6, 58.1) | --- | 45.9 (43.6, 48.2) | 41.3 (38.8, 43.7) | -15 (-18, -11) | <0.001 | --- |  |
| Overweight | 65.4 (63.5, 67.3) | --- | 56.0 (53.8, 58.2) | 48.9 (46.8, 51.0) | -16 (-19, -14) | <0.001 | -1.9 (-6.2, 2.5) | 0.398 |
| Obesity | 68.6 (65.4, 71.7) | --- | 56.6 (53.5, 59.8) | 49.0 (46.0, 52.1) | -20 (-24, -15) | <0.001 | -5.0 (-10.6, 0.7) | 0.084 |
| Class I | 70.3 (66.9, 73.7) | --- | 57.3 (53.7, 60.9) | 49.5 (45.9, 53.2) | -21 (-26, -16) | <0.001 | -6.2 (-12.3, 0.0) | 0.049 |
| Class II-III | 61.2 (54.6, 67.8) | --- | 53.9 (47.8, 59.9) | 49.3 (42.9, 55.8) | -12 (-21, -3) | 0.012 | 2.7 (-7.1, 12.6) | 0.583 |

*Notes:* Estimates age-standardised to the pooled HSE data; - - - comparable data not available. Normal weight: BMI 18.5 to <25kg/m<sup>2</sup>; overweight: BMI 25 to <30kg/m<sup>2</sup>; obesity: BMI ≥30kg/m<sup>2</sup>; Class I obesity: BMI 30 to <34.9kg/m<sup>2</sup>; Class II-III obesity: BMI ≥35kg/m<sup>2</sup>.
Estimates for total diabetes are shown in Table 5. *P*-values obtained using Wald tests.

**TABLE 3.** Age-standardised risk factor prevalence by BMI group and four-year survey period in women.

| **Risk factor by BMI group** | **2003-06** | **2007-10** | **2011-14** | **2015-18** | **PP change** | **P-value** | **PP change versus normal-weight** | **P-value** |
| --- | --- | --- | --- | --- | --- | --- | --- | --- |
|  | **% (95% CI)** | **% (95% CI)** | **% (95% CI)** | **% (95% CI)** | **% (95% CI)** |  | **% (95% CI)** |  |
| **Current smoking** |  |  |  |  |  |  |  |  |
| All | 23.4 (22.7, 24.2) | 20.1 (19.4, 20.8) | 17.8 (17.1, 18.5) | 15.6 (14.9, 16.3) | -7.8 (-8.9, -6.8) | <0.001 | --- | --- |
| Normal-weight | 25.1 (24.0, 26.2) | 20.8 (19.7, 21.9) | 18.1 (17.0, 19.2) | 16.0 (14.9, 17.2) | -9.1 (-10.7, -7.5) | <0.001 | --- | --- |
| Overweight | 22.5 (21.3, 23.7) | 20.0 (18.8, 21.2) | 17.8 (16.6, 19.0) | 15.3 (14.1, 16.5) | -7.2 (-8.9, -5.5) | <0.001 | 1.9 (-0.4, 4.1) | 0.099 |
| Obesity | 22.9 (21.4, 24.3) | 20.8 (19.3, 22.2) | 18.0 (16.6, 19.4) | 15.6 (14.3, 16.8) | -7.3 (-9.2, -5.4) | <0.001 | 1.7 (-0.7, 4.2) | 0.162 |
| Class I | 23.3 (21.5, 25.2) | 20.1 (18.3, 21.9) | 17.6 (15.8, 19.3) | 15.6 (13.9, 17.2) | -7.8 (-10.2, -5.3) | <0.001 | 1.3 (-1.6, 4.2) | 0.375 |
| Class II-III | 22.3 (20.0, 24.6) | 21.9 (19.6, 24.2) | 18.6 (16.5, 20.8) | 15.3 (13.4, 17.3) | -7.0 (-10.0, -3.9) | <0.001 | 2.1 (-1.3, 5.5) | 0.223 |
| **Inactivity** |  |  |  |  |  |  |  |  |
| All | --- | 23.6 (22.5, 24.6) | 24.4 (22.8, 25.9) | 23.2 (21.6, 24.8) | -0.4 (-2.3, 1.5) | 0.710 | --- | --- |
| Normal-weight | --- | 20.5 (19.0, 22.0) | 19.3 (17.0, 21.6) | 17.7 (15.7, 19.7) | -2.8 (-5.3, -0.3) | 0.027 | --- | --- |
| Overweight | --- | 20.2 (18.5, 21.8) | 23.3 (20.7, 25.8) | 23.2 (20.3, 26.0) | 3.0 (-0.4, 6.3) | 0.080 | 5.8 (1.8, 9.8) | 0.004 |
| Obesity | --- | 31.5 (29.1, 33.8) | 31.3 (28.0, 34.5) | 31.2 (27.6, 34.7) | -0.3 (-4.5, 3.9) | 0.887 | 2.5 (-2.3, 7.3) | 0.308 |
| Class I | --- | 28.5 (25.8, 31.3) | 30.3 (26.1, 34.5) | 28.7 (24.2, 33.1) | 0.1 (-5.1, 5.3) | 0.960 | 2.9 (-2.7, 8.6) | 0.309 |
| Class II-III | --- | 36.7 (32.6, 40.8) | 33.1 (28.3, 37.9) | 34.6 (29.3, 39.9) | -2.1 (-8.8, 4.6) | 0.539 | 0.7 (-6.4, 7.8) | 0.844 |
| **Alcohol above daily limits** |  |  |  |  |  |  |  |  |
| All | --- | 32.4 (31.5, 33.2) | 28.6 (27.7, 29.4) | 27.7 (26.8, 28.6) | -4.7 (-5.9, -3.5) | <0.001 | --- | --- |
| Normal-weight | --- | 34.3 (33.1, 35.5) | 30.5 (29.2, 31.7) | 30.7 (29.4, 32.0) | -3.5 (-5.3, -1.8) | <0.001 | --- | --- |
| Overweight | --- | 34.0 (32.5, 35.4) | 30.3 (28.9, 31.8) | 29.2 (27.7, 30.7) | -4.8 (-6.9, -2.7) | <0.001 | -1.2 (-3.9, 1.4) | 0.361 |
| Obesity | --- | 27.6 (26.0, 29.1) | 23.8 (22.3, 25.3) | 23.1 (21.6, 24.6) | -4.5 (-6.7, -2.4) | <0.001 | -1.0 (-3.7, 1.8) | 0.491 |
| Class I | --- | 27.9 (26.0, 29.8) | 25.3 (23.4, 27.3) | 24.3 (22.4, 26.3) | -3.5 (-6.2, -0.8) | 0.010 | 0.0 (-3.2, 3.2) | 0.998 |
| Class II-III | --- | 27.3 (24.8, 29.7) | 21.4 (19.1, 23.7) | 21.3 (19.0, 23.5) | -6.0 (-9.3, -2.7) | <0.001 | -2.5 (-6.2, 1.3) | 0.201 |
| **Hypertension** |  |  |  |  |  |  |  |  |
| All | 27.1 (26.4, 27.8) | 27.8 (27.0, 28.5) | 25.7 (24.9, 26.4) | 24.2 (23.4, 25.0) | -2.9 (-4.0, -1.8) | <0.001 | --- | --- |
| Normal-weight | 20.9 (19.7, 22.0) | 19.7 (18.5, 20.8) | 18.3 (17.1, 19.4) | 16.5 (15.4, 17.6) | -4.3 (-5.9, -2.7) | <0.001 | --- | --- |
| Overweight | 26.3 (25.1, 27.4) | 26.2 (25.0, 27.4) | 24.7 (23.5, 25.9) | 23.2 (21.9, 24.5) | -3.1 (-4.8, -1.4) | <0.001 | 1.2 (-1.1, 3.6) | 0.305 |
| Obesity | 37.6 (35.9, 39.3) | 40.7 (39.0, 42.5) | 36.6 (35.0, 38.2) | 34.3 (32.5, 36.1) | -3.3 (-5.8, -0.8) | 0.009 | 1.0 (-1.9, 4.0) | 0.491 |
| Class I | 34.3 (32.3, 36.2) | 35.5 (33.6, 37.4) | 34.2 (32.3, 36.2) | 30.0 (27.9, 32.0) | -4.3 (-7.1, -1.5) | 0.003 | 0.0 (-3.2, 3.3) | 0.987 |
| Class II-III | 43.4 (40.4, 46.3) | 50.3 (47.0, 53.6) | 40.5 (37.7, 43.3) | 40.7 (37.6, 43.9) | -2.6 (-7.0, 1.7) | 0.232 | 1.7 (-2.9, 6.3) | 0.471 |
| **Raised total cholesterol** |  |  |  |  |  |  |  |  |
| All | 64.7 (63.6, 65.7) | --- | 52.5 (51.3, 53.6) | 48.2 (47.0, 49.4) | -16 (-18, -15) | <0.001 | --- | --- |
| Normal-weight | 62.3 (60.7, 63.9) | --- | 51.1 (49.4, 52.8) | 46.2 (44.5, 47.9) | -16 (-18, -14) | <0.001 | --- | --- |
| Overweight | 66.0 (63.9, 68.0) | --- | 54.2 (52.3, 56.1) | 50.9 (48.4, 53.3) | -15 (-18, -12) | <0.001 | 0.9 (-3.0, 4.9) | 0.635 |
| Obesity | 71.1 (68.6, 73.7) | --- | 55.8 (53.1, 58.5) | 50.1 (47.4, 52.7) | -21 (-25, -17) | <0.001 | -5.0 (-9.3, -0.6) | 0.025 |
| Class I | 72.0 (68.9, 75.0) | --- | 57.7 (54.3, 61.2) | 50.0 (47.0, 53.0) | -22 (-26, -18) | <0.001 | -5.8 (-10.8, -0.9) | 0.021 |
| Class II-III | 69.8 (65.5, 74.0) | --- | 52.3 (48.2, 56.5) | 49.4 (45.1, 53.8) | -20 (-26, -14) | <0.001 | -4.3 (-10.8, 2.3) | 0.200 |

*Notes:* Estimates age-standardised to the pooled HSE data; - - - comparable data not available. Normal weight: BMI 18.5 to <25kg/m<sup>2</sup>; overweight: BMI 25 to <30kg/m<sup>2</sup>; obesity: BMI ≥30kg/m<sup>2</sup>; Class I obesity: BMI 30 to <34.9kg/m<sup>2</sup>; Class II-III obesity: BMI ≥35kg/m<sup>2</sup>.
Estimates for total diabetes are shown in Table 5. *P*-values obtained using Wald tests.

**Figure 1.**
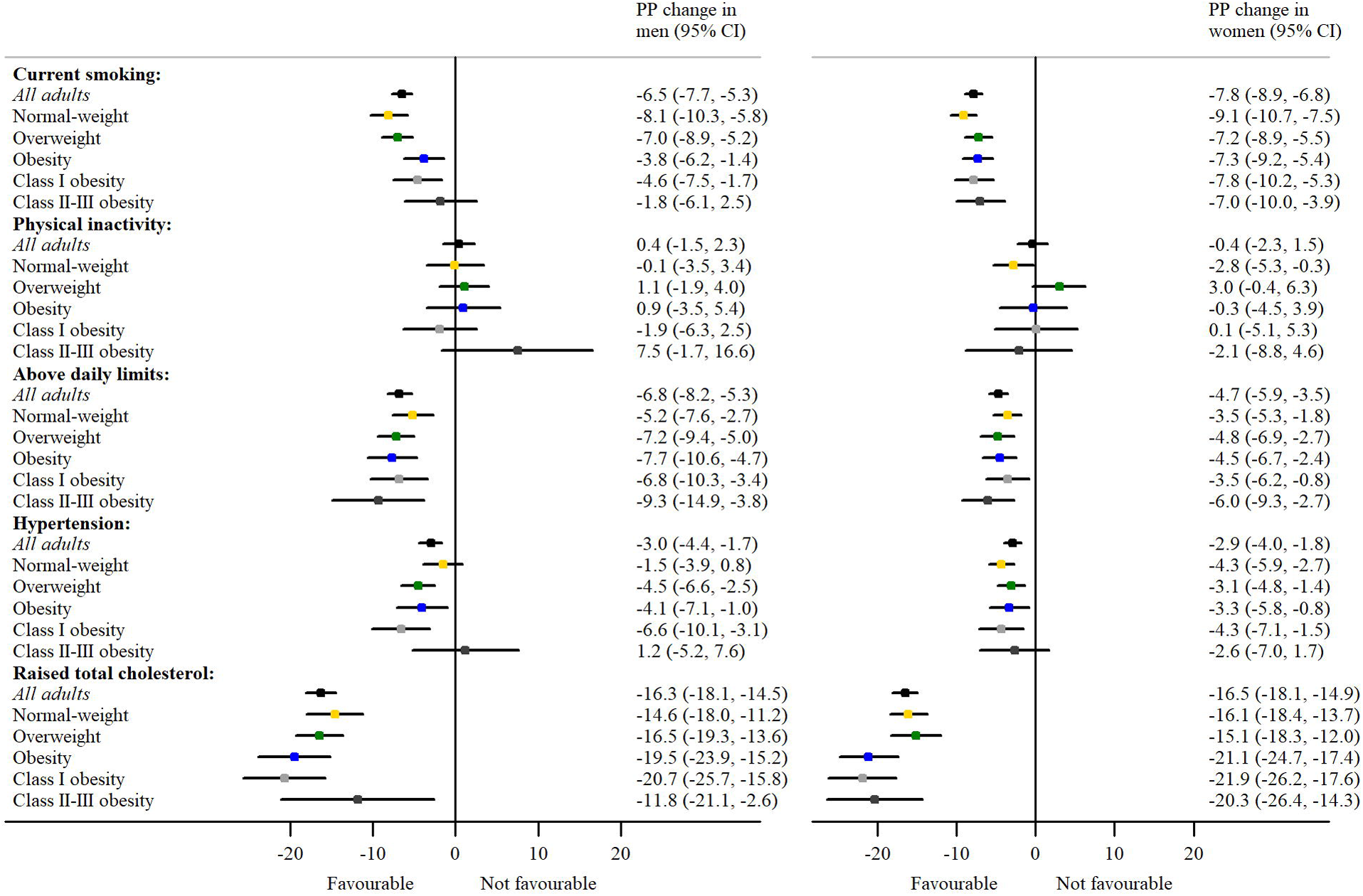

### Current cigarette smoking

Current cigarette smoking prevalence decreased among all adults in both genders from 2003–06 to 2015–18 (men: –6.5 PP; 95% CI: –7.7, –5.3; women: –7.8 PP; 95% CI: –8.9, –6.8). Smoking prevalence varied by BMI category within each time-period among men (highest among those with normal-weight) but showed no variation among women. Whilst current cigarette smoking prevalence decreased between the first and last time-periods among all BMI groups, it fell more sharply for men with normal-weight (−8.1 PP; 95% CI: –10.3, –5.8) versus men with obesity (−3.8 PP; 95% CI: –6.2, –1.4) (Figure 1).

### Physical inactivity

Levels of physical inactivity (< 30 minutes MVPA per week) were higher among adults with obesity versus those with normal-weight in each survey (2008, 2012, 2016), especially among women. Inactivity prevalence remained at a stable level among all adults (∼18% and ∼24% for men and women, respectively). However, stability in prevalence among all adults masked divergent trends by BMI category among women: inactivity prevalence decreased among those with normal-weight (−2.8 PP; 95% CI: –5.3, –0.3) but marginally increased among those overweight (3.0 PP; 95% CI: –0.4, 6.3).

### Above daily limits of alcohol consumption

Among all adults, levels of drinking above recommended daily alcohol limits decreased in both genders from 2007–10 to 2015–18 (men: –6.8 PP; 95% CI: –8.2, –5.3; women: –4.7 PP; 95% CI: –5.9, –3.5). In each time-period, levels of drinking above recommended daily limits in men were higher in the overweight than normal-weight group; levels of drinking above recommended daily limits in women were higher among those with normal-weight versus those with obesity. Prevalence decreased over time among all BMI groups for both genders (Figure 1) but change over time (relative to those with normal-weight) did not vary by BMI *(p* > 0.170).

### Survey-defined hypertension and indicators of management

Hypertension prevalence (BP ≥140/90mmHg or taking medication prescribed for high BP) decreased among all adults from 2003–06 to 2015–18 (men: –3.0 PP; 95% CI: –4.4, –1.7; women: –2.9 PP; 95% CI: –4.0, –1.8). Hypertension prevalence showed a strong graded association in both genders in each time-period, being highest among adults with obesity. Hypertension prevalence decreased among all BMI groups for both genders (by 3 to 4 PP) with the exception of no statistically significant change among men with normal-weight (−1.5 PP; 95% CI: –3.9, 0.8; *p* = 0.199).

Amongst participants with hypertension, levels of diagnosed, treated and controlled hypertension by survey period and BMI category are shown in Table 4; Figure 2 shows the change in prevalence between the first and last time-periods. The pattern of change (relative to those with normal-weight) was similar by BMI for both genders *(p* > 0.100): the proportion of hypertension that was diagnosed remained at a similar level, whilst proportions of hypertension that were treated and controlled improved during the study period.

**TABLE 4.** Levels of diagnosed, treated and controlled hypertension by BMI group, gender and four-year survey period.

| **Hypertension management indicator by BMI category** | **2003-06** | **2007-10** | **2011-14** | **2015-18** | **PP change** | **P-value** | **PP change versus normal-weight** | **P-value** |
| --- | --- | --- | --- | --- | --- | --- | --- | --- |
|  | **% (95% CI)** | **% (95% CI)** | **% (95% CI)** | **% (95% CI)** | **% (95% CI)** |  | **% (95% CI)** |  |
| **Men** |  |  |  |  |  |  |  |  |
| **Diagnosed** |  |  |  |  |  |  |  |  |
| All | 55.3 (53.1, 57.6) | 57.0 (54.1, 59.9) | 58.0 (55.8, 60.1) | 59.0 (56.9, 61.0) | 3.6 (0.6, 6.6) | 0.018 | --- | --- |
| Normal-weight | 46.3 (40.5, 52.0) | 39.8 (32.3, 47.3) | 51.1 (45.6, 56.7) | 49.3 (44.1, 54.6) | 3.1 (-4.7, 10.9) | 0.437 | --- | --- |
| Overweight | 55.4 (52.3, 58.4) | 56.1 (51.8, 60.4) | 54.8 (51.5, 58.1) | 57.7 (54.5, 60.9) | 2.3 (-2.1, 6.8) | 0.300 | -0.7 (-9.7, 8.2) | 0.871 |
| Obesity | 60.5 (56.7, 64.2) | 65.3 (60.8, 69.8) | 63.6 (60.3, 66.9) | 63.8 (60.8, 66.9) | 3.4 (-1.4, 8.2) | 0.169 | 0.3 (-8.9, 9.5) | 0.950 |
| Class I | 57.6 (53.5, 61.8) | 63.3 (58.2, 68.5) | 61.7 (57.6, 65.7) | 62.0 (58.2, 65.8) | 4.4 (-1.3, 10.0) | 0.131 | 1.3 (-8.4, 11.0) | 0.798 |
| Class II-III | 69.9 (62.6, 77.3) | 74.0 (66.0, 82.0) | 67.5 (61.8, 73.2) | 67.3 (62.2, 72.4) | -2.6 (-11.6, 6.3) | 0.561 | -5.7 (-17.5, 6.0) | 0.339 |
| **Treated** |  |  |  |  |  |  |  |  |
| All | 40.0 (38.4, 41.7) | 46.9 (45.1, 48.6) | 48.8 (46.9, 50.6) | 50.4 (48.5, 52.4) | 10.4 (7.9, 13.0) | <0.001 | --- | --- |
| Normal-weight | 33.6 (29.6, 37.7) | 39.9 (35.5, 44.3) | 40.1 (35.3, 44.9) | 41.2 (36.2, 46.2) | 7.6 (1.1, 14.0) | 0.021 | --- | --- |
| Overweight | 38.5 (36.2, 40.8) | 44.7 (42.2, 47.2) | 45.6 (43.0, 48.3) | 47.3 (44.2, 50.4) | 8.8 (4.9, 12.7) | <0.001 | 1.2 (-6.4, 8.8) | 0.755 |
| Obesity | 45.6 (42.8, 48.3) | 51.8 (49.1, 54.6) | 55.6 (52.8, 58.5) | 56.9 (54.0, 59.8) | 11.3 (7.4, 15.3) | <0.001 | 3.7 (-3.8, 11.3) | 0.329 |
| Class I | 43.0 (40.0, 46.1) | 49.6 (46.5, 52.8) | 53.2 (49.8, 56.6) | 52.4 (48.9, 55.9) | 9.4 (4.8, 14.0) | <0.001 | 1.8 (-6.2, 9.8) | 0.658 |
| Class II-III | 53.6 (47.8, 59.4) | 57.7 (52.3, 63.1) | 61.3 (56.5, 66.0) | 65.3 (60.9, 69.8) | 11.7 (4.5, 19.0) | 0.002 | 4.1 (-5.4, 13.7) | 0.394 |
| **Controlled** |  |  |  |  |  |  |  |  |
| All | 20.0 (18.6, 21.4) | 27.4 (25.8, 29.0) | 30.3 (28.6, 32.1) | 32.9 (31.0, 34.8) | 12.9 (10.6, 15.3) | <0.001 | --- | --- |
| Normal-weight | 15.9 (12.4, 19.4) | 24.6 (20.7, 28.6) | 25.9 (21.6, 30.3) | 27.0 (22.4, 31.6) | 11.1 (5.3, 16.8) | <0.001 | --- | --- |
| Overweight | 19.0 (17.0, 20.9) | 26.6 (24.3, 28.9) | 28.4 (25.9, 30.9) | 30.3 (27.4, 33.1) | 11.3 (7.8, 14.8) | <0.001 | 0.2 (-6.6, 7.0) | 0.955 |
| Obesity | 23.3 (20.8, 25.8) | 29.3 (26.7, 31.9) | 34.3 (31.4, 37.1) | 37.4 (34.6, 40.3) | 14.1 (10.3, 17.9) | <0.001 | 3.1 (-3.9, 10.0) | 0.390 |
| Class I | 22.6 (19.9, 25.4) | 28.0 (25.0, 31.0) | 33.1 (29.7, 36.4) | 34.4 (31.0, 37.7) | 11.8 (7.4, 16.1) | <0.001 | 0.7 (-6.6, 8.0) | 0.855 |
| Class II-III | 26.3 (20.4, 32.2) | 32.7 (27.3, 38.1) | 37.2 (31.9, 42.5) | 43.0 (37.7, 48.3) | 16.7 (8.8, 24.6) | <0.001 | 5.6 (-4.2, 15.4) | 0.262 |
| **Women** |  |  |  |  |  |  |  |  |
| **Diagnosed** |  |  |  |  |  |  |  |  |
| All | 60.5 (58.2, 62.9) | 65.3 (62.3, 68.3) | 65.9 (63.8, 68.0) | 63.5 (61.5, 65.4) | 3.0 (-0.1, 6.0) | 0.056 | --- | --- |
| Normal-weight | 53.3 (48.6, 58.1) | 56.1 (49.7, 62.5) | 57.1 (52.3, 62.0) | 57.5 (53.2, 61.9) | 4.2 (-2.2, 10.6) | 0.202 | --- | --- |
| Overweight | 58.4 (54.8, 62.1) | 62.3 (57.2, 67.4) | 66.6 (63.0, 70.2) | 61.1 (57.6, 64.5) | 2.6 (-2.5, 7.7) | 0.313 | -1.6 (-9.6, 6.5) | 0.702 |
| Obesity | 68.3 (64.8, 71.8) | 72.3 (68.2, 76.4) | 71.0 (68.0, 74.0) | 68.5 (65.6, 71.4) | 0.2 (-4.4, 4.7) | 0.936 | -4.0 (-11.7, 3.8) | 0.313 |
| Class I | 66.3 (61.9, 70.8) | 71.9 (66.8, 77.1) | 70.3 (66.5, 74.1) | 67.5 (63.6, 71.3) | 1.1 (-4.7, 7.0) | 0.704 | -3.0 (-11.7, 5.6) | 0.493 |
| Class II-III | 73.2 (67.4, 79.0) | 72.1 (65.4, 78.8) | 72.8 (68.2, 77.5) | 69.4 (65.0, 73.8) | -3.8 (-11.1, 3.5) | 0.307 | -8.0 (-17.5, 1.6) | 0.102 |
| **Treated** |  |  |  |  |  |  |  |  |
| All | 49.9 (48.1, 51.8) | 56.9 (55.1, 58.7) | 58.8 (57.1, 60.6) | 58.2 (56.3, 60.1) | 8.2 (5.6, 10.9) | <0.001 | --- | --- |
| Normal-weight | 40.5 (36.9, 44.0) | 46.6 (42.9, 50.3) | 51.2 (47.3, 55.2) | 49.4 (45.1, 53.6) | 8.9 (3.4, 14.4) | 0.002 | --- | --- |
| Overweight | 47.9 (45.0, 50.8) | 54.6 (51.6, 57.6) | 59.4 (56.3, 62.4) | 57.1 (53.7, 60.6) | 9.2 (4.7, 13.8) | <0.001 | 0.4 (-6.7, 7.5) | 0.918 |
| Obesity | 58.4 (55.6, 61.2) | 64.5 (62.0, 67.1) | 63.6 (61.0, 66.2) | 64.1 (61.4, 66.9) | 5.8 (1.8, 9.7) | 0.004 | -3.1 (-9.8, 3.6) | 0.363 |
| Class I | 56.1 (52.6, 59.6) | 61.9 (58.5, 65.2) | 62.1 (58.7, 65.5) | 60.2 (56.5, 63.9) | 4.1 (-1.0, 9.2) | 0.116 | -4.8 (-12.3, 2.7) | 0.210 |
| Class II-III | 63.2 (58.4, 67.9) | 68.5 (64.4, 72.6) | 66.5 (62.5, 70.5) | 68.2 (64.0, 72.4) | 5.0 (-1.3, 11.4) | 0.122 | -3.8 (-12.1, 4.4) | 0.361 |
| **Controlled** |  |  |  |  |  |  |  |  |
| All | 24.9 (23.3, 26.4) | 31.7 (29.9, 33.4) | 36.6 (34.8, 38.5) | 35.8 (33.9, 37.7) | 10.9 (8.5, 13.4) | <0.001 | --- | --- |
| Normal-weight | 20.0 (17.0, 23.0) | 27.1 (23.7, 30.5) | 30.4 (26.6, 34.2) | 32.4 (28.3, 36.5) | 12.4 (7.4, 17.5) | <0.001 | --- | --- |
| Overweight | 23.9 (21.5, 26.4) | 30.6 (27.8, 33.4) | 38.6 (35.4, 41.7) | 35.3 (32.0, 38.6) | 11.3 (7.2, 15.5) | <0.001 | -1.1 (-7.6, 5.4) | 0.744 |
| Obesity | 28.9 (26.2, 31.6) | 35.2 (32.5, 38.0) | 39.4 (36.5, 42.3) | 38.3 (35.3, 41.3) | 9.4 (5.4, 13.4) | <0.001 | -3.0 (-9.5, 3.4) | 0.361 |
| Class I | 28.2 (25.0, 31.3) | 32.1 (28.6, 35.5) | 38.6 (35.0, 42.2) | 35.6 (31.6, 39.5) | 7.4 (2.4, 12.4) | 0.004 | -5.0 (-12.2, 2.1) | 0.167 |
| Class II-III | 31.1 (26.1, 36.2) | 39.7 (35.0, 44.4) | 40.9 (35.9, 45.9) | 41.4 (36.7, 46.1) | 10.3 (3.4, 17.2) | 0.004 | -2.1 (-10.7, 6.5) | 0.627 |

*Notes:* Hypertension defined as BP $\geq 140/90$ mmHg or reported taking medication prescribed for high BP. Estimates age-standardised to the pooled HSE data (persons with hypertension). Normal weight: BMI 18.5 to $<25$ kg/m<sup>2</sup>; overweight: BMI 25 to $<30$ kg/m<sup>2</sup>; obesity: BMI $\geq 30$ kg/m<sup>2</sup>; Class I obesity: BMI 30 to $<34.9$ kg/m<sup>2</sup>; Class II-III obesity: BMI $\geq 35$ kg/m<sup>2</sup>. *P*-values obtained using Wald tests.

**Figure 2.**
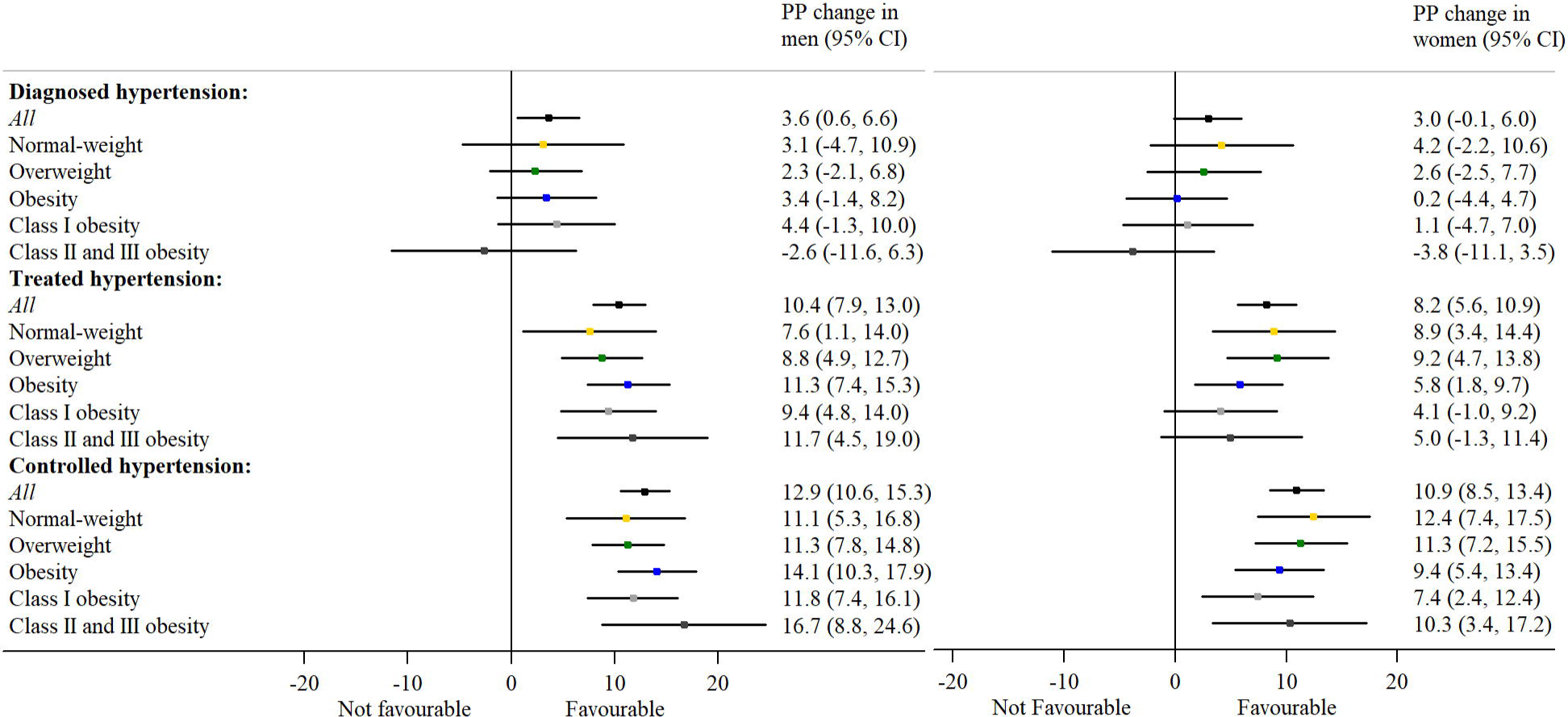

### Total diabetes, including diagnosed and undiagnosed

Estimates of total diabetes, including diagnosed and undiagnosed (elevated HbA_1c_), are shown in Table 5 (Figure 3 shows the change in prevalence between the first and last time-periods). Total diabetes increased among all adults from 2003–06 to 2015–18 (men: 2.3 PP; 95% CI: 1.3, 3.2; women: 2.0 PP; 95% CI: 1.3, 2.7). As with hypertension, total diabetes showed a strong graded association in both genders in each time-period, being highest among adults with obesity. The secular increase in total diabetes among all adults masked divergent trends by BMI category. Among men, total diabetes prevalence remained stable among adults with normal-weight (1.1 PP; 95% CI: –0.4, 2.6) but increased among adults with overweight (1.5 PP; 95% CI: 0.2, 2.7) and obesity (3.5 PP; 95% CI: 1.2, 5.7). A similar finding was observed for women.

Based on all adults as the denominator, levels of diagnosed diabetes increased among adults with overweight (men: 1.2 PP; 95% CI: 0.1, 2.3; women: 1.7 PP; 95% CI: 0.6, 2.7) and among women with obesity (2.6 PP; 95% CI: 1.0, 4.2). Levels of undiagnosed diabetes increased only among adults with obesity (men: 1.8 PP; 95% CI: 0.2, 3.4; women: 1.0 PP; 95% CI: 0.1, 1.9). Relative to their counterparts with normal-weight, levels of diagnosed diabetes increased among adults with overweight and obesity in women, but not in men.

**TABLE 5.** Levels of total diabetes, including diagnosed and undiagnosed diabetes, by BMI group, gender and four-year survey period.

| Diabetes by BMI category | 2003-06 | 2007-10 | 2011-14 | 2015-18 | PP change | P-value | PP change versus normal-weight | P-value |
| --- | --- | --- | --- | --- | --- | --- | --- | --- |
|  | % (95% CI) | % (95% CI) | % (95% CI) | % (95% CI) | % (95% CI) |  | % (95% CI) |  |
| **Men** |  |  |  |  |  |  |  |  |
| **Total diabetes** |  |  |  |  |  |  |  |  |
| All | 5.9 (5.3, 6.5) | 7.4 (6.4, 8.4) | 8.0 (7.4, 8.7) | 8.1 (7.4, 8.9) | 2.3 (1.3, 3.2) | <0.001 | --- | --- |
| Normal-weight | 3.1 (2.2, 4.0) | 2.7 (1.4, 4.0) | 4.4 (3.4, 5.4) | 4.2 (2.9, 5.4) | 1.1 (-0.4, 2.6) | 0.158 | --- | --- |
| Overweight | 5.1 (4.2, 5.9) | 6.1 (4.9, 7.4) | 5.7 (4.9, 6.5) | 6.5 (5.6, 7.5) | 1.5 (0.2, 2.7) | 0.023 | 0.4 (-1.5, 2.2) | 0.700 |
| Obesity | 10.4 (8.9, 11.9) | 12.8 (10.5, 15.0) | 14.9 (13.2, 16.6) | 13.9 (12.2, 15.6) | 3.5 (1.2, 5.7) | 0.003 | 2.4 (-0.2, 5.0) | 0.076 |
| Class I | 8.4 (6.9, 10.0) | 11.2 (8.7, 13.8) | 12.3 (10.4, 14.2) | 11.4 (9.6, 13.3) | 3.0 (0.6, 5.5) | 0.015 | 1.9 (-0.8, 4.7) | 0.170 |
| Class II-III | 17.5 (13.4, 21.5) | 18.3 (13.2, 23.5) | 22.6 (18.9, 26.4) | 20.4 (16.7, 24.1) | 3.0 (-2.5, 8.4) | 0.291 | 1.8 (-3.8, 7.5) | 0.524 |
| **Diagnosed** |  |  |  |  |  |  |  |  |
| All | 4.2 (3.7, 4.7) | 5.5 (4.6, 6.3) | 5.9 (5.3, 6.4) | 5.9 (5.3, 6.5) | 1.7 (0.9, 2.5) | <0.001 | --- | --- |
| Normal-weight | 2.3 (1.5, 3.1) | 2.2 (1.0, 3.3) | 3.7 (2.8, 4.6) | 3.7 (2.4, 4.9) | 1.4 (-0.1, 2.8) | 0.063 | --- | --- |
| Overweight | 3.6 (2.8, 4.4) | 4.4 (3.3, 5.4) | 4.2 (3.6, 4.9) | 4.8 (4.0, 5.6) | 1.2 (0.1, 2.3) | 0.039 | -0.2 (-2.0, 1.6) | 0.825 |
| Obesity | 7.5 (6.1, 8.8) | 9.3 (7.4, 11.3) | 10.3 (8.9, 11.6) | 9.2 (8.0, 10.3) | 1.7 (-0.1, 3.5) | 0.062 | 0.3 (-2.0, 2.6) | 0.790 |
| Class I | 5.7 (4.4, 7.1) | 8.0 (5.9, 10.1) | 8.3 (6.8, 9.8) | 7.5 (6.3, 8.8) | 1.8 (0.0, 3.7) | 0.053 | 0.4 (-1.9, 2.8) | 0.713 |
| Class II-III | 14.0 (10.2, 17.7) | 14.2 (9.5, 18.9) | 16.3 (13.3, 19.4) | 13.6 (10.9, 16.4) | -0.3 (-4.9, 4.3) | 0.899 | -1.7 (-6.5, 3.2) | 0.497 |
| **Undiagnosed** |  |  |  |  |  |  |  |  |
| All | 1.7 (1.3, 2.0) | 2.0 (1.4, 2.5) | 2.1 (1.8, 2.5) | 2.2 (1.9, 2.6) | 0.6 (0.1, 1.1) | 0.021 | --- | --- |
| Normal-weight | 0.8 (0.3, 1.3) | 0.6 (0.0, 1.2) | 0.7 (0.4, 1.1) | 0.5 (0.2, 0.9) | -0.3 (-0.8, 0.3) | 0.350 | --- | --- |
| Overweight | 1.5 (1.1, 1.9) | 1.8 (1.0, 2.5) | 1.5 (1.1, 1.9) | 1.8 (1.2, 2.3) | 0.3 (-0.4, 1.0) | 0.383 | 0.6 (-0.3, 1.4) | 0.183 |
| Obesity | 2.9 (2.1, 3.8) | 3.4 (2.1, 4.8) | 4.6 (3.5, 5.8) | 4.7 (3.4, 6.0) | 1.8 (0.2, 3.4) | 0.029 | 2.0 (0.3, 3.7) | 0.019 |
| Class I | 2.7 (1.8, 3.6) | 3.3 (1.7, 4.8) | 4.0 (2.7, 5.4) | 3.9 (2.4, 5.4) | 1.2 (-0.5, 2.9) | 0.174 | 1.5 (-0.4, 3.3) | 0.115 |
| Class II-III | 3.5 (1.5, 5.5) | 4.2 (1.3, 7.0) | 6.3 (3.7, 8.8) | 6.8 (3.9, 9.6) | 3.2 (-0.2, 6.7) | 0.067 | 3.5 (0.0, 7.1) | 0.050 |
| **Women** |  |  |  |  |  |  |  |  |
| **Total diabetes** |  |  |  |  |  |  |  |  |
| All | 4.0 (3.5, 4.5) | 5.4 (4.6, 6.3) | 5.9 (5.4, 6.5) | 6.0 (5.4, 6.5) | 2.0 (1.3, 2.7) | <0.001 | --- | --- |
| Normal-weight | 2.6 (1.9, 3.3) | 2.0 (1.2, 2.9) | 2.2 (1.7, 2.8) | 2.4 (1.8, 2.9) | -0.2 (-1.1, 0.7) | 0.648 | --- | --- |
| Overweight | 3.3 (2.7, 4.0) | 4.1 (2.9, 5.2) | 4.8 (4.0, 5.5) | 5.2 (4.3, 6.2) | 1.9 (0.8, 3.1) | 0.001 | 2.1 (0.7, 3.6) | 0.004 |
| Obesity | 7.3 (6.0, 8.6) | 11.6 (8.9, 14.3) | 12.0 (10.4, 13.5) | 10.9 (9.6, 12.2) | 3.6 (1.8, 5.4) | <0.001 | 3.8 (1.8, 5.8) | <0.001 |
| Class I | 5.8 (4.5, 7.1) | 7.4 (5.4, 9.4) | 10.1 (8.5, 11.8) | 8.4 (6.9, 9.8) | 2.5 (0.6, 4.5) | 0.010 | 2.8 (0.7, 4.8) | 0.010 |
| Class II-III | 9.8 (7.2, 12.5) | 17.8 (12.7, 23.0) | 15.3 (12.2, 18.4) | 14.9 (12.6, 17.1) | 5.0 (1.5, 8.5) | 0.005 | 5.2 (1.6, 8.8) | 0.004 |
| **Diagnosed** |  |  |  |  |  |  |  |  |
| All | 3.1 (2.6, 3.5) | 3.6 (3.0, 4.3) | 4.1 (3.6, 4.6) | 4.5 (4.0, 5.0) | 1.5 (0.8, 2.1) | <0.001 | --- | --- |
| Normal-weight | 2.1 (1.4, 2.7) | 1.2 (0.6, 1.9) | 1.2 (0.8, 1.6) | 1.7 (1.2, 2.2) | -0.4 (-1.2, 0.4) | 0.367 | --- | --- |
| Overweight | 2.5 (1.9, 3.1) | 2.5 (1.7, 3.4) | 3.3 (2.7, 4.0) | 4.2 (3.3, 5.1) | 1.7 (0.6, 2.7) | 0.002 | 2.1 (0.8, 3.4) | 0.002 |
| Obesity | 5.5 (4.3, 6.6) | 7.6 (5.9, 9.3) | 8.5 (7.2, 9.9) | 8.1 (7.0, 9.2) | 2.6 (1.0, 4.2) | 0.001 | 3.0 (1.2, 4.7) | 0.001 |
| Class I | 4.2 (3.1, 5.3) | 5.6 (3.8, 7.4) | 6.9 (5.5, 8.4) | 5.7 (4.5, 7.0) | 1.5 (-0.1, 3.2) | 0.073 | 1.9 (0.1, 3.7) | 0.041 |
| Class II-III | 7.7 (5.3, 10.1) | 10.6 (7.5, 13.7) | 11.4 (8.6, 14.3) | 11.8 (9.7, 13.9) | 4.2 (1.0, 7.3) | 0.010 | 4.5 (1.3, 7.8) | 0.006 |
| **Undiagnosed** |  |  |  |  |  |  |  |  |
| All | 0.9 (0.7, 1.2) | 1.8 (1.3, 2.3) | 1.8 (1.5, 2.1) | 1.5 (1.2, 1.7) | 0.5 (0.2, 0.9) | 0.002 | --- | --- |
| Normal-weight | 0.5 (0.2, 0.8) | 0.8 (0.3, 1.3) | 1.0 (0.7, 1.4) | 0.7 (0.3, 1.0) | 0.2 (-0.3, 0.6) | 0.505 | --- | --- |
| Overweight | 0.8 (0.5, 1.1) | 1.5 (0.8, 2.3) | 1.4 (1.0, 1.8) | 1.1 (0.7, 1.4) | 0.2 (-0.2, 0.7) | 0.332 | 0.1 (-0.6, 0.8) | 0.811 |
| Obesity | 1.8 (1.2, 2.5) | 4.0 (1.8, 6.2) | 3.4 (2.6, 4.2) | 2.8 (2.2, 3.5) | 1.0 (0.1, 1.9) | 0.025 | 0.9 (-0.2, 1.9) | 0.097 |
| Class I | 1.6 (0.9, 2.4) | 1.8 (0.8, 2.8) | 3.2 (2.3, 4.2) | 2.6 (1.8, 3.4) | 1.0 (-0.1, 2.1) | 0.068 | 0.8 (-0.3, 2.0) | 0.160 |
| Class II-III | 2.2 (0.9, 3.4) | 7.2 (2.8, 11.6) | 3.9 (2.5, 5.3) | 3.0 (2.0, 4.0) | 0.9 (-0.7, 2.5) | 0.285 | 0.7 (-1.0, 2.4) | 0.403 |

Notes: Undiagnosed diabetes: no reported diagnosed diabetes and HbA<sub>1c</sub> ≥6.5% (prior to HSE 2012) or ≥48mmol/mol (HSE 2012-18). Estimates age-standardised to the pooled HSE data. Normal weight: BMI 18.5 to <25kg/m<sup>2</sup>; overweight: BMI 25 to <30kg/m<sup>2</sup>; obesity: BMI ≥30kg/m<sup>2</sup>; Class I obesity: BMI 30 to <34.9kg/m<sup>2</sup>; Class II-III obesity: BMI ≥35kg/m<sup>2</sup>. *P*-values obtained using Wald tests.

**Figure 3.**
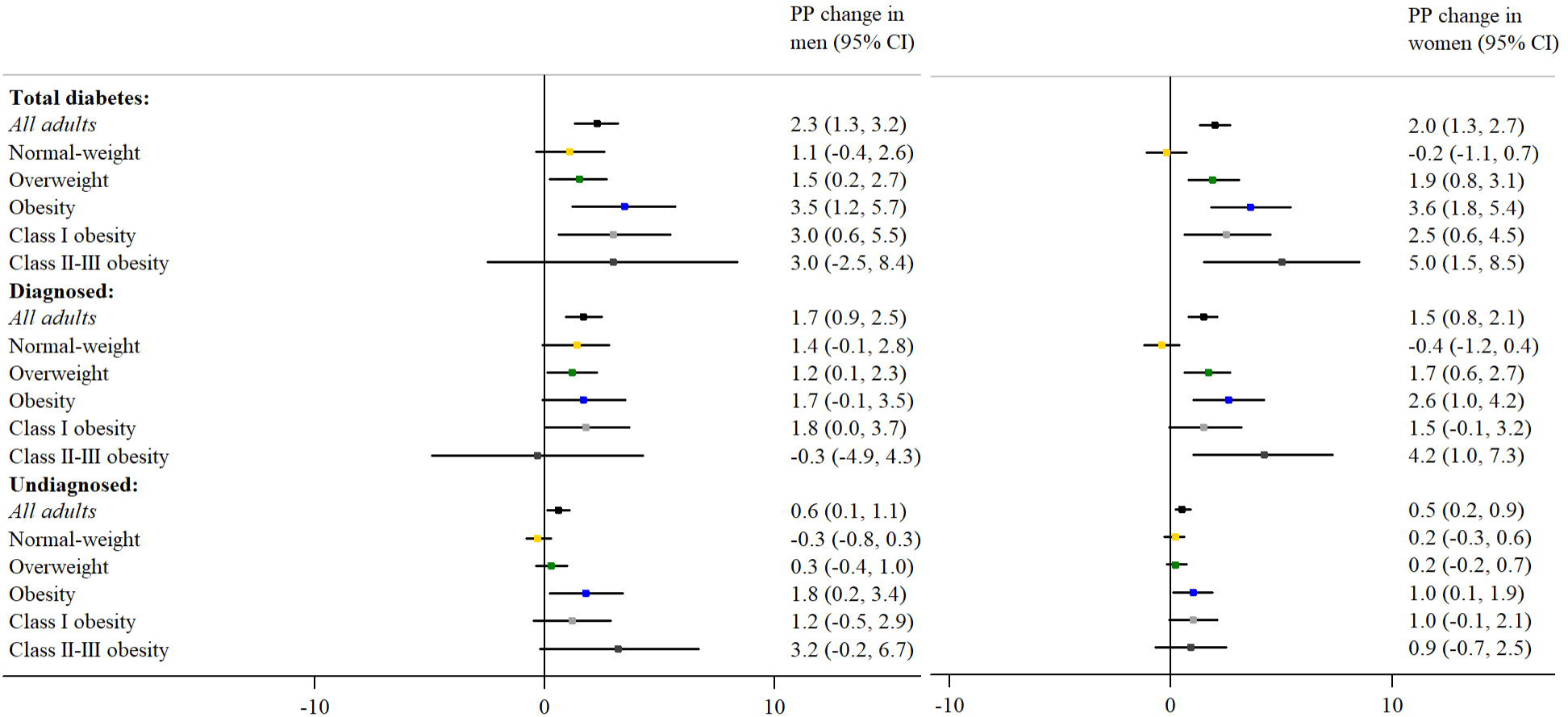

### Raised total cholesterol

Levels of raised total cholesterol (≥5mmol/L) decreased among all adults from 2003–06 to 2015–18 (men: –16 PP; 95% CI: –18, –14; women: –16 PP; 95% CI: –18, –15). Cross-sectionally, raised total cholesterol prevalence was highest among adults with overweight and obesity. Raised total cholesterol prevalence decreased among all BMI groups, but fell more sharply among women with obesity (−21 PP; 95% CI: –25, –17) versus their counterparts with normal-weight (−16 PP; 95% CI: –18, –14).

## Discussion

Using data spanning 16 years (2003–18), we examined change over time in the prevalence of six key CVD risk factors by BMI category. Whilst levels of physical inactivity and consumption of alcohol above daily limits (on the heaviest drinking day) were stable and decreased in all BMI groups respectively, a number of risk factors showed divergent trends. First, whilst current cigarette smoking prevalence decreased among all BMI groups, it declined more slowly among men with obesity. Secondly, hypertension prevalence decreased among all BMI groups for both genders, except among men with normal-weight. Thirdly, among both genders, total diabetes prevalence remained stable among normal-weight adults, but increased among adults with overweight and obesity. Fourthly, raised total cholesterol prevalence decreased among all BMI groups for both genders, but fell more sharply among women with obesity.

## Comparisons with other studies

Among adults with overweight and obesity, our findings of: (i) decreases in hypertension and raised cholesterol, and (ii) increases in diabetes, agree with similar analyses of the US National Health and Nutrition Examination Survey (NHANES). Among adults aged 20–74 years with overweight and obesity, Gregg and colleagues using 1960–2000 data found (i) decreasing levels of high BP and current smoking, (ii) a stable level of total diabetes, and an(iii) increase in diagnosed diabetes (7). Increases in treated hypertension were larger among adults with overweight and obesity than for lean (BMI < 25kg/m^2^) adults (7). Among adults with obesity, Saydah and colleagues using 1999–2010 data found: (i) stable levels of self-reported smoking, total diabetes, undiagnosed diabetes, and hypertension, and (ii) a decrease in untreated hypertension (6). Guo and Garvey using 1999–2014 data reported a significant increase in mean HbA_1c_ among adults with obesity, whereas mean BP and lipid metrics improved (18).

## Population-level trends in CVD risk factor prevalence

In agreement with the population-level trends presented here, previous studies using HSE data have shown decreases in current cigarette smoking (19), hypertension (19), raised total cholesterol (19); stability in PA (13, 20); and increases in total diabetes (19) and in obesity (5). Multiple policies have been enacted in England over the study period which have likely affected these trends. These included attempts to standardise and improve the management of chronic diseases in primary care settings, e.g. financially incentivised screening and treatment of hypertension and dyslipidaemia with lifestyle advice and/or medications (21) and published guidelines by the National Institute for Health and Care Excellence (NICE) which recommended targeted screening to identify undiagnosed diabetes in asymptomatic populations (22). National health promotional activities included a widely marketed mass media “This girl can” programme to increase PA (23). Voluntary targets for industry were set by governments, such as reducing the salt content of processed foods (24). Whole-population based strategies included tobacco control policies such as smoke-free legislation, higher taxation, a higher minimum age of sale of cigarettes, licensing of nicotine replacement therapy for harm reduction, and introduction of plain packaging (25).

Despite these policies, the population-level trends in CVD risk factors show both favourable and unfavourable change. The decline in current cigarette smoking prevalence has been linked to the tobacco control policies listed above (26). The recent fall in survey-defined hypertension is typically attributed to decreased salt intake in foods (24) and improved detection, treatment and control of high BP, although levels of management remain suboptimal (27). Whilst levels of glycaemic control have improved across all social groups (28), the rise in total diabetes reflects both improved case ascertainment and increases in incidence associated with rising obesity (through numerous pathophysiological mechanisms that increase the risk of type II diabetes among adults with obesity) (29). Longer survival for those with diabetes is also a potential contributory factor (30). The increased uptake and efficacy of lipid-lowering medications such as statins for the primary prevention of CVD within UK primary care during the study period (31) may be a key driver of the reduction in raised total cholesterol prevalence.

## Divergent trends in CVD risk factor prevalence by BMI category

A number of factors have likely contributed to the divergent trends in risk factors by BMI category. Larger reductions in the prevalence of survey-defined hypertension and raised total cholesterol among adults with overweight and obesity may reflect at least partially the results of the aforementioned targeted efforts to improve chronic disease management in primary care settings (6, 7). In terms of CVD risk profile, diagnosing existing cases of diabetes is generally beneficial, particularly for BP and lipid modification as well as glycaemic control, resulting in a stalling or reduction of risk factor progression despite advancing disease (15). Worldwide reductions in raised levels of BP and cholesterol through improved treatment and/or changes in other risks have contributed to the fall in CVD rates worldwide, despite increases in BMI (3).

In contrast, the slower fall in cigarette smoking prevalence among men with obesity, albeit from a lower prevalence in the first survey period (2003–06), and the marginal increase in inactivity prevalence among women with overweight, may to some extent reflect influences of living in obesogenic environments (32): i.e. neighbourhoods with higher socioeconomic deprivation, geographic barriers to PA, and lower air quality that may influence adiposity levels over and above other individual characteristics (33). Reverse causality, i.e. a reluctance of smokers to tolerate post-cessation weight gain (34), and that higher adiposity in itself is a risk-factor for smoking (35), making it more difficult to quit, is also a potential contributory factor for the slower decline in current smoking levels among men with obesity.

## Strengths and limitations

The main strengths of our study include the use of repeated nationally-representative health examination surveys that objectively measured anthropometry, BP, cholesterol and HbA_1c_ using standardised protocols over the 16-year study-period, thereby eliminating self-report bias for these factors (36). Stratifying results by gender enabled us to identify differences in the patterns of change over time; our use of direct standardisation removed the potential confounding influence of age.

The present study has a number of limitations. Our study is descriptive and so does not directly address the underlying causes of the recent divergent trends in CVD risk factors by BMI. Use of repeated cross-sectional surveys with new samples drawn annually precludes assessment of within-individual change in BMI or risk factors. However, this design provides the ability to measure undiagnosed disease, which would be unethical in longitudinal studies. Data on smoking, alcohol, PA and diagnosed diabetes relied on self-reported information and may be subject to recall and social desirability bias. Despite the pooling of annual data to improve precision, limitations of sample size meant that we could not examine trends in risk factors by BMI within different minority ethnic groups, or examine trends in diagnosed diabetes amongst those with total diabetes. Response rates to the HSE have declined over time, creating the potential for increased bias in the most recent survey years, although the overall survey response rate, in and of itself, is not a good indicator of the level of non-response bias (37). In the present study, participants who were interviewed but excluded from the analytical sample due to missing anthropometry data were significantly older, less educated, and less likely to report very good/good general health; this proportion has also increased over time, from 12% in 2003 to 18% in 2018 (data not shown). We used non-response weights available with the data to minimise the impact of response bias on our findings; nevertheless, our findings may underestimate the differences in risk factor prevalence by BMI and overestimate the magnitude of favourable trends. Finally, changes in clinical guidelines for reducing high levels of BP, total cholesterol and HbA_1c_ can make the long-term interpretation of trends difficult as adults would be more likely to be treated at lower levels of CVD risk in the most recent surveys (7). However, the elevated levels of BP (≥140/90mmHg) (14), total cholesterol (≥5mmol/L) and HbA_1c_ (≥48mmol/mol) (38) used in our study were based on guidelines relevant for the duration of the study period.

## Conclusion

Relative to normal-weight adults, greater reductions in hypertension and raised total cholesterol among adults with overweight and obesity reflect at least in part improvements in screening and treatment in those at highest cardiovascular risk. Higher levels of risk factor prevalence among adults with overweight and obesity in parallel with secular increases in diabetes highlight the importance of national prevention efforts to combat the public health impact of excess adiposity.

## Data Availability

The HSE datasets generated and analysed during the current study are available via the UK Data Service (UKDS: https://ukdataservice.ac.uk/), subject to their end user license agreement. Statistical code to enable replication of our results (using the datasets deposited at the UKDS) is available on request from the corresponding author. Citations for the HSE datasets are provided at the end of this manuscript (this is below):
University College London, Department of Epidemiology and Public Health, National Centre for Social Research. (2010). Health Survey for England, 2003. [data collection]. 2nd Edition. UK Data Service. SN: 5098, http://doi.org/10.5255/UKDA-SN-5098-1.
National Centre for Social Research, University College London. Department of Epidemiology and Public Health. (2011). Health Survey for England, 2005. [data collection]. 3rd Edition. UK Data Service. SN: 5675, http://doi.org/10.5255/UKDA-SN-5675-1.
National Centre for Social Research, University College London. Department of Epidemiology and Public Health. (2011). Health Survey for England, 2006. [data collection]. 4th Edition. UK Data Service. SN: 5809, http://doi.org/10.5255/UKDA-SN-5809-1.
National Centre for Social Research, University College London. Department of Epidemiology and Public Health. (2010). Health Survey for England, 2007. [data collection]. 2nd Edition. UK Data Service. SN: 6112, http://doi.org/10.5255/UKDA-SN-6112-1.
National Centre for Social Research, University College London. Department of Epidemiology and Public Health. (2013). Health Survey for England, 2008. [data collection]. 4th Edition. UK Data Service. SN: 6397, http://doi.org/10.5255/UKDA-SN-6397-2.
National Centre for Social Research, University College London. Department of Epidemiology and Public Health. (2015). Health Survey for England, 2009. [data collection]. 3rd Edition. UK Data Service. SN: 6732, http://doi.org/10.5255/UKDA-SN-6732-2.
NatCen Social Research, Royal Free and University College Medical School. Department of Epidemiology and Public Health. (2015). Health Survey for England, 2010. [data collection]. 3rd Edition. UK Data Service. SN: 6986, http://doi.org/10.5255/UKDA-SN-6986-3.
NatCen Social Research, University College London. Department of Epidemiology and Public Health. (2013). Health Survey for England, 2011. [data collection]. UK Data Service. SN: 7260, http://doi.org/10.5255/UKDA-SN-7260-1.
NatCen Social Research, University College London. Department of Epidemiology and Public Health. (2014). Health Survey for England, 2012. [data collection]. UK Data Service. SN: 7480, http://doi.org/10.5255/UKDA-SN-7480-1.
NatCen Social Research, University College London. Department of Epidemiology and Public Health. (2015). Health Survey for England, 2013. [data collection]. UK Data Service. SN: 7649, http://doi.org/10.5255/UKDA-SN-7649-1.
NatCen Social Research, University College London. Department of Epidemiology and Public Health. (2016). Health Survey for England, 2014. [data collection]. 2nd Edition. UK Data Service. SN: 7919, http://doi.org/10.5255/UKDA-SN-7919-2.
NatCen Social Research, University College London. Department of Epidemiology and Public Health. (2017). Health Survey for England, 2015. [data collection]. UK Data Service. SN: 8280, http://doi.org/10.5255/UKDA-SN-8280-1.
NatCen Social Research, University College London. Department of Epidemiology and Public Health. (2019). Health Survey for England, 2016. [data collection]. 3rd Edition. UK Data Service. SN: 8334, http://doi.org/10.5255/UKDA-SN-8334-3.
NatCen Social Research, University College London. Department of Epidemiology and Public Health. (2020). Health Survey for England, 2017. [data collection]. 2nd Edition. UK Data Service. SN: 8448, http://doi.org/10.5255/UKDA-SN-8448-2.
NatCen Social Research, University College London. Department of Epidemiology and Public Health. (2020). Health Survey for England, 2018. [data collection]. 2nd Edition. UK Data Service. SN: 8649, http://doi.org/10.5255/UKDA-SN-8649-1.

## Availability of data and materials

The HSE datasets generated and analysed during the current study are available via the UK Data Service (UKDS: https://ukdataservice.ac.uk/), subject to their end user license agreement. Statistical code to enable replication of our results (using the datasets deposited at the UKDS) is available on request from the corresponding author. Citations for the HSE datasets are provided at the end of this manuscript.

## Abbreviations

BMI: body mass index
BP: blood pressure
CVD: cardiovascular disease
HSE: Health Survey for England
MVPA: moderate-to-vigorous physical activity
NCDs: noncommunicable diseases
NHANES: National Health and Nutrition Examination Survey
PA: physical activity
PP: percentage change
UKDS: UK Data Service
WHO: World Health Organization

## Acknowledgements

The authors thank our colleagues at NatCen Social Research, the interviewers and nurses, and the participants in the Health Survey for England series.

## Contributions

SS and JSM conceptualized the study. SS was responsible for generating the datasets, conducting the analyses, interpreting the results and drafting the manuscript. SS, LNF and JSM critically revised the manuscript. All authors have read and approved the manuscript.

## Ethics approval

The procedures used in the HSE to obtained informed consent from survey participants are very closely scrutinised by a NHS ethics committee each year. Approval was obtained from the following Research Ethics Committees (REC): HSE 2003, 2005, 2006 and 2007: London Multi-Centre REC; HSE 2008: Oxford A REC: 07/H0604/102; HSE 2009: Oxford B REC: 08/H0605/103; HSE 2010: Oxford B REC: 09/H0605/73; HSE 2011 and 2012: Oxford A REC: 10/H0604/56; HSE 2013 and 2014: Oxford A REC: 12/sc/0317; HSE 2015: West London NRES Committee: 14/LO/0862; HSE 2016, 2017 and 2018: Nottingham REC: 15/EE/0299. This study is a secondary analysis of previously collected data and so additional ethical approval was not required.

## Consent for publication

Not applicable

## Appendix Citations for the HSE datasets

University College London, Department of Epidemiology and Public Health, National Centre for Social Research. (2010). *Health Survey for England, 2003*. [data collection]. *2nd Edition*. UK Data Service. SN: 5098, http://doi.org/10.5255/UKDA-SN-5098-1.
National Centre for Social Research, University College London. Department of Epidemiology and Public Health. (2011). *Health Survey for England, 2005*. [data collection]. *3rd Edition.* UK Data Service. SN: 5675, http://doi.org/10.5255/UKDA-SN-5675-1.
National Centre for Social Research, University College London. Department of Epidemiology and Public Health. (2011). *Health Survey for England, 2006*. [data collection]. *4th Edition*. UK Data Service. SN: 5809, http://doi.org/10.5255/UKDA-SN-5809-1.
National Centre for Social Research, University College London. Department of Epidemiology and Public Health. (2010). *Health Survey for England, 2007*. [data collection]. *2nd Edition.* UK Data Service. SN: 6112, http://doi.org/10.5255/UKDA-SN-6112-1.
National Centre for Social Research, University College London. Department of Epidemiology and Public Health. (2013). *Health Survey for England, 2008*. [data collection]. *4th Edition*. UK Data Service. SN: 6397, http://doi.org/10.5255/UKDA-SN-6397-2.
National Centre for Social Research, University College London. Department of Epidemiology and Public Health. (2015). *Health Survey for England, 2009*. [data collection]. *3rd Edition.* UK Data Service. SN: 6732, http://doi.org/10.5255/UKDA-SN-6732-2.
NatCen Social Research, Royal Free and University College Medical School. Department of Epidemiology and Public Health. (2015). *Health Survey for England, 2010*. [data collection]. 3rd Edition. UK Data Service. SN: 6986, http://doi.org/10.5255/UKDA-SN-6986-3.
NatCen Social Research, University College London. Department of Epidemiology and Public Health. (2013). *Health Survey for England, 2011*. [data collection]. UK Data Service. SN: 7260, http://doi.org/10.5255/UKDA-SN-7260-1.
NatCen Social Research, University College London. Department of Epidemiology and Public Health. (2014). *Health Survey for England, 2012*. [data collection]. UK Data Service. SN: 7480, http://doi.org/10.5255/UKDA-SN-7480-1.
NatCen Social Research, University College London. Department of Epidemiology and Public Health. (2015). *Health Survey for England, 2013*. [data collection]. UK Data Service. SN: 7649, http://doi.org/10.5255/UKDA-SN-7649-1.
NatCen Social Research, University College London. Department of Epidemiology and Public Health. (2016). *Health Survey for England, 2014*. [data collection]. 2nd Edition. UK Data Service. SN: 7919, http://doi.org/10.5255/UKDA-SN-7919-2. collection]. *3rd Edition*. UK Data Service. SN: 5675, http://doi.org/10.5255/UKDA-SN-collection]. *2nd Edition*. UK Data Service. SN: 6112, http://doi.org/10.5255/UKDA-SN-collection]. *3rd Edition*. UK Data Service. SN: 6732, http://doi.org/10.5255/UKDA-SN-
NatCen Social Research, University College London. Department of Epidemiology and Public Health. (2017). *Health Survey for England, 2015*. [data collection]. UK Data Service. SN: 8280, http://doi.org/10.5255/UKDA-SN-8280-1.
NatCen Social Research, University College London. Department of Epidemiology and Public Health. (2019). *Health Survey for England, 2016*. [data collection]. 3^rd^ Edition. UK Data Service. SN: 8334, http://doi.org/10.5255/UKDA-SN-8334-3.
NatCen Social Research, University College London. Department of Epidemiology and Public Health. (2020). *Health Survey for England, 2017*. [data collection]. 2nd Edition. UK Data Service. SN: 8448, http://doi.org/10.5255/UKDA-SN-8448-2.
NatCen Social Research, University College London. Department of Epidemiology and Public Health. (2020). *Health Survey for England, 2018*. [data collection]. 2nd Edition. UK Data Service. SN: 8649, http://doi.org/10.5255/UKDA-SN-8649-1.

## References

1. Di Angelantonio E, Bhupathiraju SN, Wormser D, Gao P, Kaptoge S, de Gonzalez AB, et al. Body-mass index and all-cause mortality: individual-participant-data meta-analysis of 239 prospective studies in four continents. The Lancet. 2016;388(10046):776–86.

2. Khan SS, Ning H, Wilkins JT, Allen N, Carnethon M, Berry JD, et al. Association of body mass index with lifetime risk of cardiovascular disease and compression of morbidity. JAMA Cardiology. 2018;3(4):280–7.

3. GBD 2015 Obesity Collaborators. Health effects of overweight and obesity in 195 countries over 25 years. New England Journal of Medicine. 2017;377(1):13–27.

4. Devaux M, Lerouge A, Ventelou B, Goryakin Y, Feigl A, Vuik S, et al. Assessing the potential outcomes of achieving the World Health Organization global non-communicable diseases targets for risk factors by 2025: is there also an economic dividend? Public health. 2019;169:173–9.

5. Conolly A, Craig S. Health Survey for England 2018. Overweight and obesity in adults and children. NHS Digital, Leeds; 2019.

6. Saydah S, Bullard KM, Cheng Y, Ali MK, Gregg EW, Geiss L, et al. Trends in cardiovascular disease risk factors by obesity level in adults in the United States, NHANES 1999 2010. Obesity. 2014;22(8):1888–95.

7. Gregg EW, Cheng YJ, Cadwell BL, Imperatore G, Williams DE, Flegal KM, et al. Secular trends in cardiovascular disease risk factors according to body mass index in US adults. JAMA. 2005;293(15):1868–74.

8. Park K, Lim S, Park Y, Ju W, Shin Y, Yeom H. Cardiovascular disease risk factors and obesity levels in Korean adults: results from the Korea National Health and Nutrition Examination Survey, 2007–2015. Osong public health and research perspectives. 2018;9(4):150.

9. Green M, Subramanian S, Razak F. Population-level trends in the distribution of body mass index in England,1992–2013. J Epidemiol Community Health. 2016;70(8):832–5.

10. Mindell J, Biddulph JP, Hirani V, Stamatakis E, Craig R, Nunn S, et al. Cohort profile: the health survey for England. International journal of epidemiology. 2012;41(6):1585–93.

11. Fuller E. Health Survey for England 2007. Healthy Lifestyles: Knowledge, Attitudes and Behaviour – Chapter 7: Adult alcohol consumption and attitudes to drinking. Leeds: The NHS Information Centre; 2008.

12. World Health Organization. World Health Organization BMI Classification. 2006.

13. Scholes S, Mindell J. Health Survey for England 2012 – Chapter 2: Physical Activity in Adults. Leeds: Health and Social Care Information Centre; 2013.

14. Mancia G, De Backer G, Dominiczak A, Cifkova R, Fagard R, Germano G, et al. 2007 Guidelines for the management of arterial hypertension: The Task Force for the Management of Arterial Hypertension of the European Society of Hypertension (ESH) and of the European Society of Cardiology (ESC). European Heart Journal. 2007;28(12):1462–536.

15. Moody A, Cowley G, Fat LN, Mindell JS. Social inequalities in prevalence of diagnosed and undiagnosed diabetes and impaired glucose regulation in participants in the Health Surveys for England series. BMJ Open. 2016;6(2):e010155.

16. National Institute for Health and Clinical Excellence. Lipid modification: cardiovascular risk assessment and the modification of blood lipids for the primary and secondary prevention of cardiovascular disease: NICE; 2008.

17. NatCen Social Research, University College London. Department of Epidemiology and Public Health. (2020). Health Survey for England, 2018. [data collection]. 2nd Edition. UK Data Service. SN: 8649, http://doi.org/10.5255/UKDA-SN-8649-1.

18. Guo F, Garvey WT. Trends in cardiovascular health metrics in obese adults: National Health and Nutrition Examination Survey (NHANES), 1988–2014. Journal of the American Heart Association. 2016;5(7):e003619.

19. Scholes S. Health Survey for England 2018. Adults’ health. NHS Digital, Leeds; 2019.

20. Scholes S, Neave A. Health Survey for England 2016 – Physical activity in Adults. Leeds: Health and Social Care Information Centre; 2017.

21. Martin Roland D. Linking physicians’ pay to the quality of care—a major experiment in the United Kingdom. N Engl J Med. 2004;351:1448–54.

22. NICE Public Health Guidance. Preventing type 2 diabetes: risk identification and interventions for individuals at high risk. NICE. 2012.

23. Sport England. This Girl Can. 2015.

24. He F, Brinsden H, MacGregor G. Salt reduction in the United Kingdom: a successful experiment in public health. Journal of human hypertension. 2014;28(6):345–52.

25. Garnett C, Tombor I, Beard E, Jackson SE, West R, Brown J. Changes in smoker characteristics in England between 2008 and 2017. Addiction. 2020;115(4):748–56.

26. Levy DT, Currie L, Clancy L. Tobacco control policy in the UK: blueprint for the rest of Europe? The European Journal of Public Health. 2013;23(2):201–6.

27. Scholes S, Conolly A, Mindell JS. Income-based inequalities in hypertension and in undiagnosed hypertension: analysis of health survey for England data. Journal of Hypertension. 2020;38(5):912–24.

28. Fleetcroft R, Asaria M, Ali S, Cookson R. Outcomes and inequalities in diabetes from 2004/2005 to 2011/2012: English longitudinal study. British Journal of General Practice. 2017;67(654):e1–e9.

29. Kahn SE, Hull RL, Utzschneider KM. Mechanisms linking obesity to insulin resistance and type 2 diabetes. Nature. 2006;444(7121):840–6.

30. Zghebi SS, Steinke DT, Carr MJ, Rutter MK, Emsley RA, Ashcroft DM. Examining trends in type 2 diabetes incidence, prevalence and mortality in the UK between 2004 and 2014. Diabetes, Obesity and Metabolism. 2017;19(11):1537–45.

31. O’Keeffe AG, Nazareth I, Petersen I. Time trends in the prescription of statins for the primary prevention of cardiovascular disease in the United Kingdom: a cohort study using The Health Improvement Network primary care data. Clinical Epidemiology. 2016;8:123.

32. Swinburn B, Egger G, Raza F. Dissecting obesogenic environments: the development and application of a framework for identifying and prioritizing environmental interventions for obesity. Preventive Medicine. 1999;29(6):563–70.

33. Davillas A, Jones AM. Regional inequalities in adiposity in England: distributional analysis of the contribution of individual-level characteristics and the small area obesogenic environment. Economics & Human Biology. 2020:100887.

34. Hasegawa K, Komiyama M, Takahashi Y. Obesity and cardiovascular risk after quitting smoking: The latest evidence. European Cardiology Review. 2019;14(1):60.

35. Carreras-Torres R, Johansson M, Haycock PC, Relton CL, Smith GD, Brennan P, et al. Role of obesity in smoking behaviour: Mendelian randomisation study in UK Biobank. BMJ. 2018;361:k1767.

36. Flegal KM, Ogden CL, Fryar C, Afful J, Klein R, Huang DT. Comparisons of Self Reported and Measured Height and Weight, BMI, and Obesity Prevalence from National Surveys: 1999–2016. Obesity. 2019;27(10):1711–9.

37. Groves RM, Peytcheva E. The impact of nonresponse rates on nonresponse bias: a meta-analysis. Public Opinion Quarterly. 2008;72(2):167–89.

38. World Health Organization. Use of glycated haemoglobin (HbA1c) in diagnosis of diabetes mellitus: abbreviated report of a WHO consultation. World Health Organization; 2011.

